# Lower respiratory tract C. albicans induces lung injury in mice and associates with worse lung injury endpoints in humans

**DOI:** 10.1101/2024.10.23.24316013

**Authors:** Nathanial J. Tolman, Wonseok Choi, Jonathan K. Alder, Mohammadreza Tabary, Shulin Qin, Xiaohong Wang, Yingze Zhang, Yizeng Tu, Lokesh Sharma, Jessica Bon, Keven Robinson, Mark Snyder, Charles Dela Cruz, Minh Hong Nguyen, Alison Morris, Partha Biswas, William Bain, Georgios D. Kitsios

**Affiliations:** Division of Pulmonary, Allergy, Critical Care and Sleep Medicine, Department of Medicine, University of Pittsburgh School of Medicine and University of Pittsburgh Medical Center, Pittsburgh, PA, USA; Department of Microbiology and Immunology, Renaissance School of Medicine, Stony Brook University, Stony Brook, NY, USA; Acute Lung Injury and Infection Center, University of Pittsburgh, Pittsburgh, PA, USA; Division of Pulmonary, Critical Care, Allergy and Immunologic Diseases, Department of Internal Medicine, Wake Forest University School of Medicine, Winston-Salem, NC, USA; Veterans Affairs Pittsburgh Healthcare System, Pittsburgh, PA, USA; Division of Infectious Diseases, Department of Medicine, University of Pittsburgh School of Medicine and University of Pittsburgh Medical Center, Pittsburgh, PA, USA

## Abstract

The recovery of *Candida* species (spp.) from lower respiratory tract (LRT) secretions in critically ill patients has traditionally been considered benign. However, emerging evidence suggests that *Candida* in the LRT may be associated with adverse clinical outcomes during mechanical ventilation. To investigate the impact of *Candida* on lung injury in mice, we performed intratracheal inoculation of *C. albicans* and assessed for lung barrier function. We found that intratracheal *C. albicans* potentiated lung barrier disruption by lipopolysaccharide. Furthermore, intratracheal *C. albicans* alone was sufficient to induce lung injury, marked by neutrophil airspace recruitment and barrier disruption. Intratracheal *C. albicans* exposure in neutrophil depleted mice (PMN^DTR^) exacerbated lung injury and led to fungal dissemination. In lung epithelial cell culture, *C. albicans* caused significant lung epithelial cytotoxicity, which was attenuated with heat-killed and yeast-locked (TNRG1) *C. albicans* strains. Human data corroborated our murine model findings, demonstrating elevated biomarkers of epithelial lung injury and worse lung injury endpoints among patients with LRT *Candida* spp. Our study challenges the dogma that LRT *Candida* is harmless, suggesting that *C. albicans* can both directly cause lung injury and exacerbate lung injury from other insults. Elucidating these host-pathogen interactions may uncover new therapeutic targets in the management of acute respiratory failure in critically ill patients.

## Introduction

*Candida* species (spp.) are among the most common organisms recovered in cultures of lower respiratory tract (LRT) secretions in critically ill patients (1, 2). Historically, the presence of *Candida* spp. in the LRT has been considered airway colonization that rarely requires antifungal treatment (3). Nonetheless, isolation of *Candida* spp. from the LRT has been associated with longer duration of mechanical ventilation and hospitalization, increased risk of ventilator-associated pneumonia (VAP), and higher mortality (4–6). Beyond culture-based studies, mycobiota profiling with DNA next-generation sequencing demonstrated that high abundance of *C. albicans* in LRT secretions from mechanically ventilated patients was independently associated with worse systemic inflammation and mortality following adjustment for clinical severity variables (7). Despite this growing body of evidence, a feasibility clinical trial with systemic antifungal treatment in *C. albicans-*colonized patients at the time of VAP diagnosis did not show results promising enough to justify a larger efficacy trial for *C. albicans* eradication with systemic antifungals (8). Thus, the clinical significance of *Candida* spp. presence in the LRT remains uncertain (9).

Animal models of LRT colonization have primarily evaluated the effect of *C. albicans* on susceptibility to secondary bacterial pneumonia with mixed results. In rat models, *C. albicans* increased susceptibility to secondary bacterial pneumonia with *A. baumanii* (10), *P. aeruginosa* (11, 12), and *S. aureus* (11), likely through impairing macrophage phagocytosis. However, in a murine model, *C. albicans* colonization protected mice from *P. aeruginosa*-induced lung injury via innate lymphoid cell recruitment and IL-22 secretion (13). Such inconsistent results may be accounted for by context-dependent interactions between *C. albicans*, the host, and each bacterial pathogen.

To overcome these challenges, we first sought to understand the impact of LRT *C. albicans* presence in the context of sterile lung injury using intratracheal instillation of lipopolysaccharide (LPS) in a murine model.. Additionally, we evaluated the effect of intratracheal *C. albicans* alone in both wild-type and neutrophil depleted mice (PMN^DTR^). We then investigated the impact of *C. albicans* on epithelial cell death and barrier disruption using *in vitro* murine and human epithelial cell lines. Finally, we analyzed clinically available data from a prospective cohort of patients with respiratory failure to determine the impact of *C. albicans* colonization on clinical outcomes. We hypothesized that LRT *C. albicans* would induce lung injury *in vivo* and associate with worse clinical outcomes in our human cohort.

## Results

### Pre-existing LRT *C. albicans* potentiates sterile lung injury in mice

We first performed dose-finding experiments to validate an intratracheal (IT) *C. albicans* (strain SC5314) murine lung colonization model, similar to prior reports (14). We found consistent recoverable *C. albicans* in lung homogenates on day 2 at a dose of 10^6^ colony forming units (CFUs) but not 10^5^ CFUs. In contrast, both doses had no difference in CFU recovery compared to vehicle by day 4 indicating clearance (Figure S1C). Finally, we confirmed that IT administration at 10^6^ CFUs did not disseminate beyond the lung compartment (Figure S1D) indicating an effective mucosal response to prevent systemic candidiasis (15). We then utilized a two-hit model with instillation of 10^6^ IT *C. albicans* CFUs followed by IT LPS when *C. albicans* is still detectable in the LRT to examine the impact of *C. albicans* presence in the LRT on sterile lung injury (Figure 1A).

**Figure 1.**
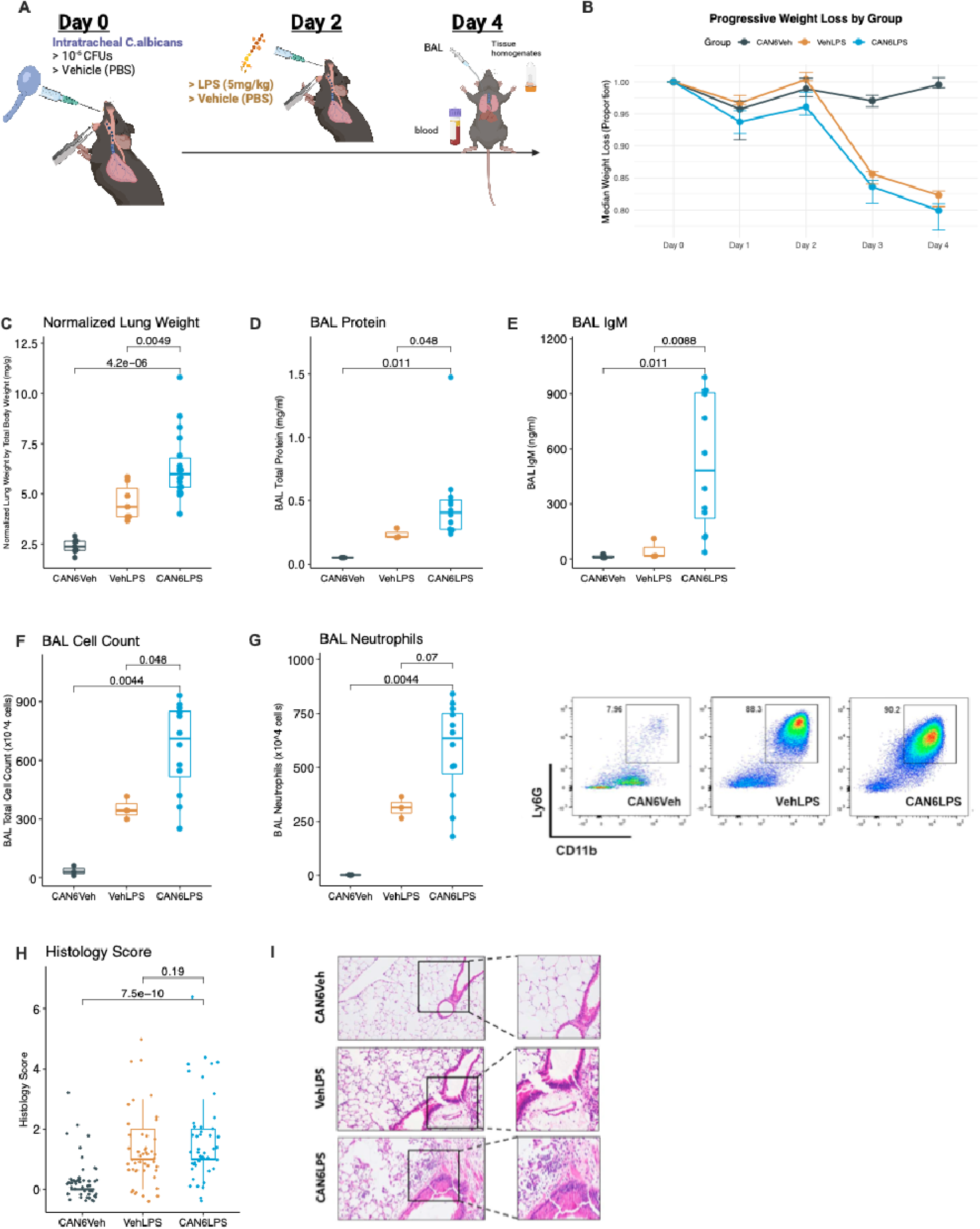
Intratracheal *C. albicans* potentiates lipopolysaccharide mediated lung injury in mice: (A) Mice received intratracheal (IT) administration of either 10^6^ colony forming units (CFUs) of *C. albicans* (CAN6) or phosphate buffered saline ([PBS], Veh) on day 0, followed by either IT lipopolysaccharide ([LPS], dosed at 5mg/kg body weight) or PBS on day 2 and were then sacrificed on day 4. Three groups were generated based on day 0 and day 2 exposures: VehLPS (n=7), CAN6LPS (n=18) and CAN6Veh (n=7). (B) Median weight loss across the experimental groups. (C) Lung weights normalized to total body weight with bronchoalveolar lavage measurements of (D) total protein by bicinchoninic acid assay and (E) IgM by enzyme-linked immunosorbent assay. (F-G) BAL total cell counts as measured by hematocytometer. Flow cytometry was used to calculate total neutrophil counts using Ly6G and CD11b, with representative scatter plots shown. (H-I) Histology scores across experimental groups graded by a blinded reviewer from 12 random fields of hematoxylin & eosin-stained lung tissue, with representative 20X image portrayed.

Wildtype C57BL6 mice were intratracheally exposed to either 10^6^ *C. albicans* (abbreviated as CAN6) or Vehicle (Veh) on day 0 followed on day 2 by either LPS (dosed at 5mg/kg) or Veh. Thus, we had three experimental groups (CAN6Veh, VehLPS, and CAN6LPS, Figure 1A) that underwent necropsy on day 4. Following a mild, transient weight loss, CAN6Veh mice regained their body weight by day 4. In contrast, mice exposed to LPS on day 2 lost further weight by day 4, with CAN6LPS mice having the numerically lowest body weight by day 4 (median 79.9%, Figure 1B). CAN6LPS mice had significantly higher normalized lung weights (by total body weight) compared to other groups (pairwise p<0.005, Figure 1C), and this difference was similar in both male and female mice (data not shown). CAN6LPS mice had significantly higher broncho-alveolar lavage (BAL) concentrations of total protein and IgM compared to both CAN6Veh and VehLPS (Figure 1D-E), indicative of worse lung barrier disruption in the presence of LRT *C. albicans* at the time of LPS insult. CAN6LPS mice had higher total BAL cell counts compared to VehLPS, and CAN6LPS and VehLPS groups had significantly more BAL neutrophilia compared to CAN6Veh mice (Figure 1F-G).

In a dedicated experiment for histologic examination, blinded histology interpretation of lung injury scores showed significantly higher lung injury scores (mean 1.50 vs. 0.0, p<0.001) in CAN6LPS vs. CAN6Veh mice, and numerically higher but not statistically different scores between CAN6LPS and VehLPS mice (median 1.50 vs. 1.00, p=0.19, Figure 1H). Representative histology slides demonstrate that the main lung injury element identified was neutrophilic infiltration into alveolar spaces (Figure 1I).

### Intratracheal *C. albicans* alone is sufficient to induce alveolar-capillary barrier disruption

We next examined whether IT *C. albicans* alone could cause lung injury, focusing on the day 2 timepoint from our 2-hit model. We therefore exposed wildtype C57BL6 mice to IT CAN6 or vehicle on Day 0 followed by necropsy on day 2 (Figure 2A). Similar to prior experiments, *C. albicans* growth was detected in both BAL and lung homogenate samples in CAN6 mice (Figure 2B), which showed mild weight loss on day 1 and 2 compared to vehicle-treated mice (Figure 2C). CAN6 mice had higher normalized lung weight as well as BAL total protein and IgM levels compared to Veh mice, suggestive of worse lung injury and barrier disruption (Figure 2D-F). CAN6 mice also had significantly higher BAL total cell counts and neutrophil counts compared to Veh mice (Figure 2G-H). Therefore, IT *C. albicans* alone was sufficient to cause lung injury and neutrophilic influx by day 2. To better understand how IT *C. albicans* causes lung injury, we next examined lung transcriptomic signatures on day 1 post IT *C. albicans*.

**Figure 2.**
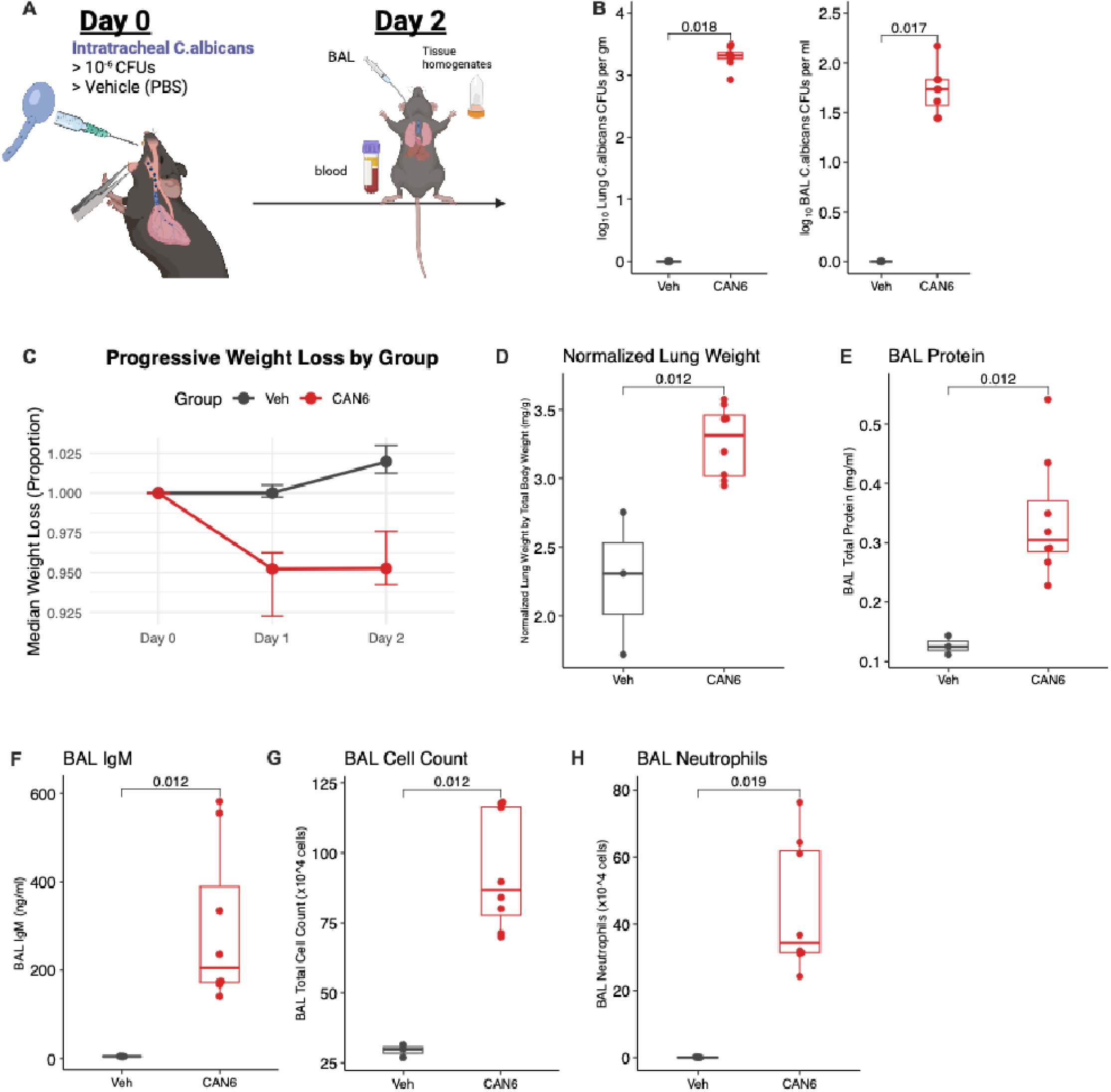
Intratracheal *C. albicans* alone is sufficient to induce alveolar-capillary barrier disruption in mice: (A) Mice were intratracheally (IT) administered 10^6^ colony forming units (CFUs) of *C. albicans* (CAN6, n=8) or phosphate-buffered saline (Veh, n=3) on day 0, followed by sacrifice on day 2. (B) Log-transformed *C. albicans* colony forming units (CFUs) per gram of lung or per milliliter of bronchoalveolar lavage fluid. (C) Median weight loss per group. (D) Lung weights normalized to total body weight with bronchoalveolar lavage measurements of (E) total protein by bicinchoninic acid assay and (F) IgM by enzyme-linked immunosorbent assay. (G) Total cell counts from bronchoalveolar lavage fluid with (I) neutrophil cell count as measured by flow cytometry.

### *C. albicans* induces a neutrophil-enriched lung transcriptome during early lung injury

We performed a dedicated experiment with wildtype C57BL6 mice exposed to IT CAN6 or vehicle on day 0 followed by necropsy on day 1 (Figure 3A). Consistent with our other experiments, CAN6 mice demonstrated increased weight loss by day 1 (Figure 3B) and lung injury marked by higher normalized lung weight, BAL total protein and IgM levels (Figure 3C-E). Additionally, soluble receptor of advanced glycation end products (sRAGE) levels, a validated biomarker of lung epithelial injury (16), were elevated in CAN6 mice (Figure 3F). CAN6 mice had markedly increased BAL total cell counts and neutrophil influx (Figure 3G,H) compared to vehicle treated mice. To further investigate the mechanisms by which *C. albicans* induced lung injury, we also performed bulk RNA sequencing on snap frozen left lungs, which had not been lavaged. Differential gene expression demonstrated up-regulation of known neutrophil activation markers (17), including *S100a8* and *S100a9* (Figure 3I) in the *C. albicans*-treated group. Furthermore, significant genes (p-adjusted < 0.05) were over-represented in biological pathways related to neutrophil migration, chemotaxis, and activation (Figure 3J). Gene set enrichment analysis using the Reactome gene list identified neutrophil degranulation and immune activation and signaling as significantly enriched in CAN6-treated lungs (Figure 3K). Given the striking phenotype of lung injury and brisk neutrophilic influx with IT *C. albicans* exposure, we hypothesized that neutrophil depletion would attenuate lung injury in our model.

**Figure 3:**
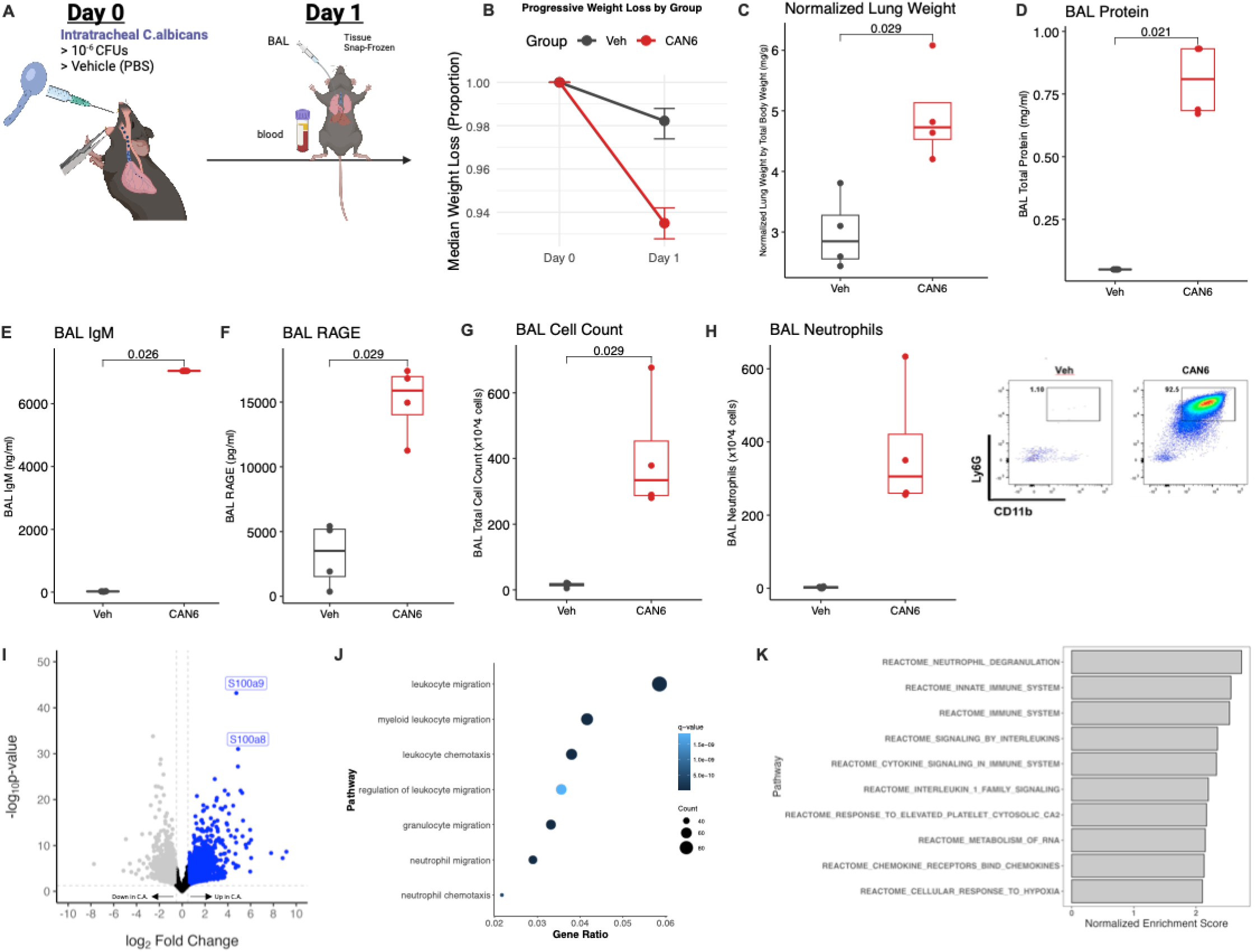
*C. albicans* induces a neutrophil-enriched lung transcriptome during early lung injury: (A) Mice received either 10^6^ colony forming units of *C. albicans* (CAN6, n=4) or phosphate buffered saline (Veh, n=4) on day 0 and were scarified on day 1. (B) Median weight loss between the experimental groups on day 1. (C) Lung weights normalized to total body weight with bronchoalveolar lavage measurements of (D) total protein by bicinchoninic acid assay and (E-F) IgM and soluble receptor for advanced glycation end products (RAGE) by enzyme-linked immunosorbent assay. (G) Bronchoalveolar lavage total cell count by hematocytometry, with (H) neutrophil cell count as measured by flow cytometry. Representative scatter plots are shown. (I-K) Transcriptomic analyses of snap frozen left lungs harvested on day 1 from mice treated with *C. albicans* compared to vehicle control. (I) Volcano plot demonstrating log_2_ fold change on the x-axis versus – log_10_(p-value) on the y-axis. Blue shading denotes up-regulation in *C. albicans* treatment, grey shading denotes down-regulation, and black is not significant. (I) Dot plot of positively regulated and over-represented genes (p-adjusted < 0.05) in *C. albicans*-treated mice. (J) Gene set enrichment analysis using the Reactome gene set with top 10 pathways plotted against their normalized enrichment score.

### Neutrophils are indispensable for clearing *C. albicans* colonization

We utilized PMN^DTR^ mice that carry both MRP8-Cre^+^ and diphtheria toxin receptor (DTR) transgenic mice (n=4) or Cre negative littermate controls (PMN^WT^, n=4). Both groups were administered intraperitoneal diphtheria toxin (500ng/mouse) 24 hours prior to inoculation and at the time of inoculation (D0), as previously described (Figure 4A) (18). Strikingly, only 50% of PMN^DTR^ mice survived by day 2 post IT *C. albicans*, whereas all PMN^WT^ mice survived by day 2 (Figure 4B). Surviving PMN^DTR^ mice continued to lose body weight by day 2, whereas PMN^WT^ mice showed partial recovery of weights (Figure 4C). Importantly, PMN^DTR^ had markedly higher fungal burden in both lung tissue homogenates and BAL (Figure 4D-E) as well as evidence of systemic *C. albicans* dissemination with high fungal burden in kidney homogenates (Figure 4F). As expected, PMN^DTR^ mice showed fewer cells in BAL (Figure 4G), with markedly reduced neutrophil counts (Figure 4H). Despite markedly reduced recruitment of neutrophils to the airspaces, surviving PMN^DTR^ mice had increased normalized lung weight, BAL levels of total protein, IgM, and sRAGE (Figure 4I-L), indicating significantly worse lung injury and barrier disruption compared to PMN^WT^ mice. Overall, these results suggest that neutrophils play essential roles in protection of the lung mucosal barrier against *C. albicans* dissemination (15, 19, 20) and limiting fungal growth and injury in the LRT. Furthermore, because lung barrier disruption was increased with neutrophil ablation, we then postulated that *C. albicans* can cause direct lung epithelial injury independently of the immune cell response.

**Figure 4.**
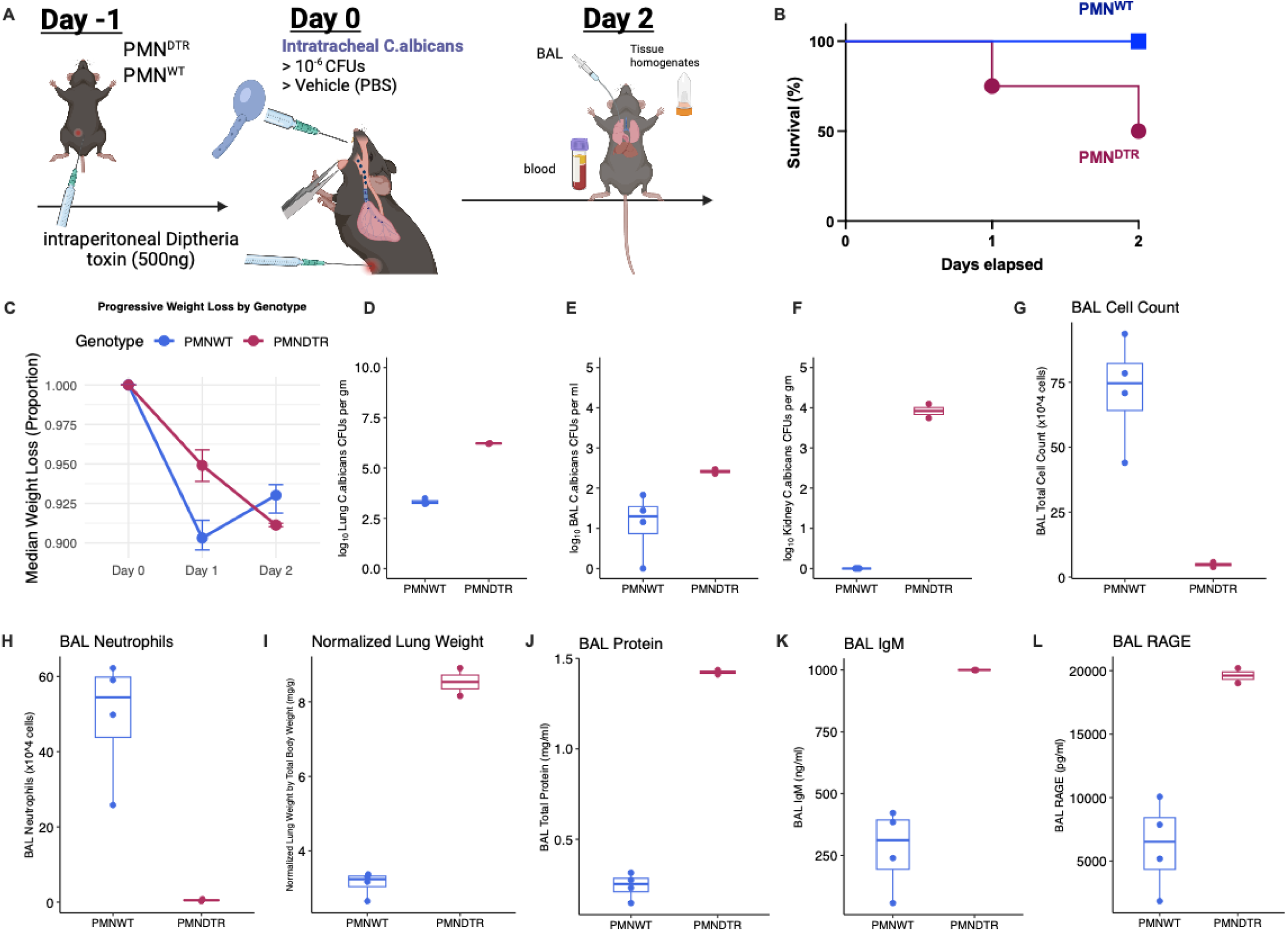
Neutrophils are indispensable for host defense against intratracheal *C. albicans*: (A) Neutrophil ablated mice (PMN^DTR^, n=4) were created by crossing MRP8-Cre^+^ mice with diphtheria toxin receptor (DTR) transgenic mice and treated with two consecutive injections of diphtheria toxin (500ng/mouse, Day –1 and 0 prior to intratracheal *C. albicans*). Littermate MRP8-Cre^-^ and DTR crossed mice (PMN^WT^, n=4) received the same diphtheria toxin injections and were used as control. (B) Kaplan-Meier survival curve for PMN^DTR^ and PMN^WT^ mice by day 2. (C) Median weight loss across experimental groups. Log-transformed colony forming units of *C. albicans* recovered on day 2 in (D) lung homogenates, (E) bronchoalveolar lavage fluid (BAL), and (F) kidney homogenates. (G) Total BAL cell count by hematocytometer and (H) neutrophil count as measured by flow cytometry. (I) Lung weights normalized to total body weight with bronchoalveolar lavage measurements of (J) total protein by bicinchoninic acid assay and (K-L) IgM and RAGE by enzyme-linked immunosorbent assay.

### *C. albicans* causes lung epithelial cytotoxicity in cell culture models

We exposed the murine lung epithelial cell line (MLE-12) to increasing multiplicities of infection (MOIs) of *C. albicans* (strain SC5314). We found that *C. albicans* administration led to a dose-dependent increase in lung epithelial cytotoxicity at 6 hours as measured by elevated LDH release in culture media (Figure 5A). We confirmed that the cytotoxic response induced by *C. albicans* required live pathogens as heat-killing *C. albicans* (HKCA) prior to administration abrogated LDH release in our murine lung epithelial cell line (Figure 5B). To investigate the mechanism(s) by which *C. albicans* induced lung epithelial cytotoxicity, we performed bulk RNA sequencing on cultured MLE-12 cells 4 hours after treatment with *C. albicans* (CA) or HKCA. Previous work in human oral epithelial cells has shown that *C. albicans* induces an innate immune response through nuclear factor-kappa-light-chain-enhancer of activated B cells (NFκB), which leads to mitogen-activated protein kinase (MAPK) signaling through morphology-independent Jun proto-oncogene (*JUN*) activation and hyphae-dependent Fos proto-oncogene (*FOS*)/dual-specificity phosphatase-1 (*DUSP1*) activation (21). Our bulk RNA sequencing data demonstrated strong upregulation of both arms, with *Junb*, *Fos*, *Fosb*, and *Dusp1* all among the most highly upregulated genes in CA-treated lung epithelial cells (Figure 5C). Gene set enrichment analysis with the Hallmark gene set identified NFκB and multiple MAPK-associated gene sets, such as p53 and KRAS, as significantly enriched (Figure 5D). Given the importance of the hyphal form in oral epithelial injury (22), we hypothesized that hyphal, but not yeast, forms would be cytotoxic to lung epithelial cells. We confirmed *C. albicans* hyphal forms abut lung epithelial cells in culture by methenamine silver staining (Figure 5E). Therefore, we next used a yeast-locked *C. albicans* (TNRG1) strain that overexpresses the negative filamentous regulator *Nrg1* gene (23) and an isogenic control (TT21) in cytotoxicity assays in MLE-12 cells. We found that the yeast-locked strain had a marked reduction in cytotoxicity compared to the isogenic control (Figure 5F). We then used a human alveolar epithelial cell line (CI-huArlo) to validate the observed cytotoxic effect of *Candida* on lung epithelial cells. Co-culture of *Candida* with human lung epithelial cells reproduced a dose-dependent cytotoxicity as measured by LDH release (Figure 5G). To investigate the impact of *C. albicans* on lung barrier integrity, we measured transepithelial resistance (TEER) using an air-liquid interface culture of human lung epithelial cells. *Candida* significantly reduced TEER at 24 hours compared to vehicle control (Figure 5H), indicating direct epithelial barrier disruption. Intriguingly, yeast-locked (TNRG1) mutant *C. albicans* did not cause a reduction in TEER compared to wild-type (TT21) control (Figure 5I). This suggests that hyphae may be required for *C. albicans*-induced lung epithelial injury and barrier disruption.

**Figure 5:**
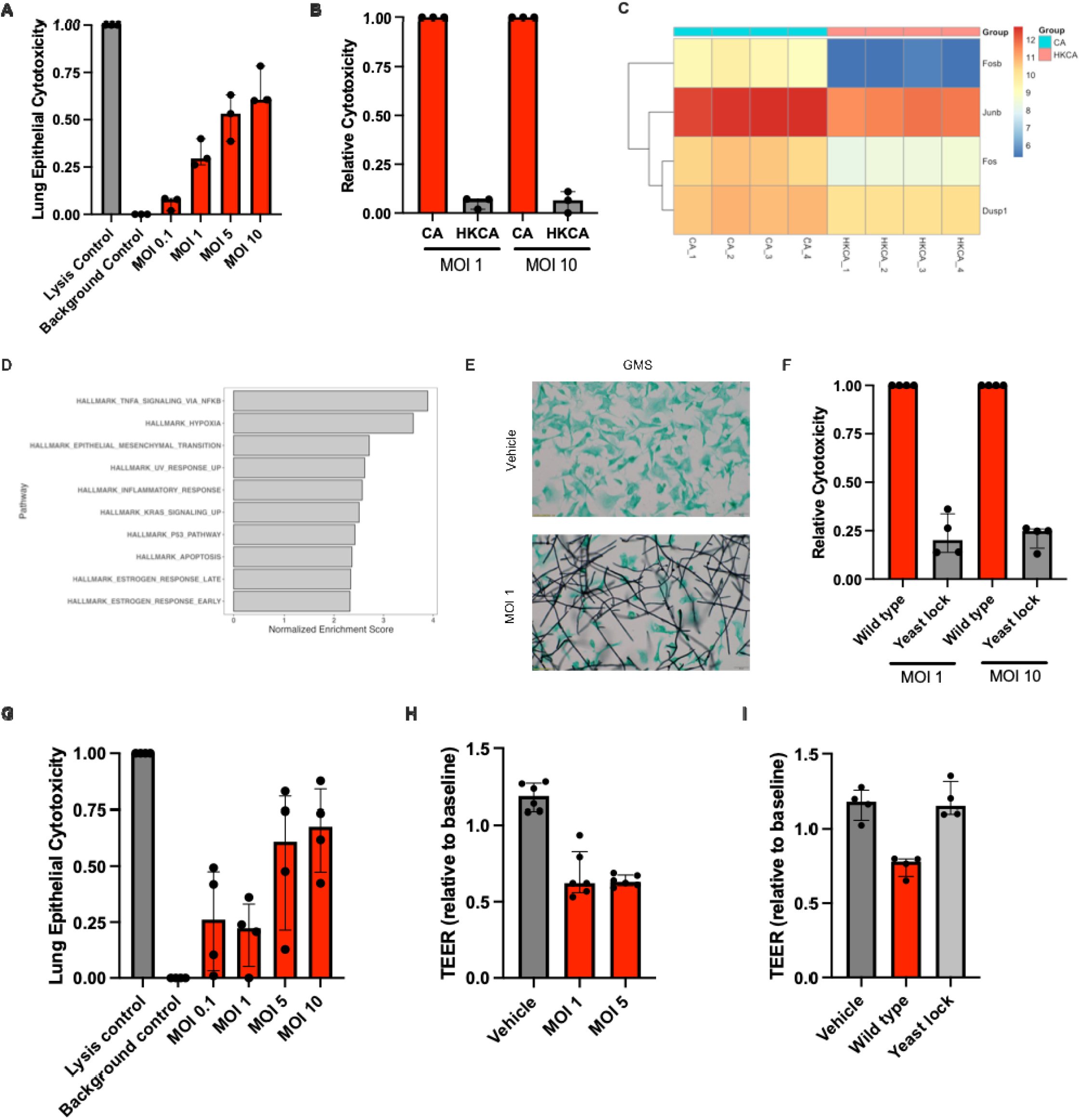
*C. albicans* causes lung epithelial cell death in murine lung epithelial cells: (A) Murine lung epithelial cell cytotoxicity as measured by lactate dehydrogenase release with *C. albicans* co-culture at differing multiplicities of infection (MOI). Values are reported as percent cytotoxicity relative to treatment with a lysis control (2% Triton-X) after subtraction of background luminescence. (B) Relative cytotoxicity of *C. albicans* to heat-killed control at MOI 1 and MOI 10. (C-D) Bulk RNA sequencing of cultured MLE-12 cells treated with *C. albicans* (CA) or heat-killed *C. albicans* (HKCA) at MOI 0.5. (C) Heatmap of log_2_ transformed genes of interest between CA and HKCA. (D) Gene set enrichment analysis of the top 10 hallmark pathways plotted against their normalized enrichment score. (E) Representative 60X images of Grocott’s methenamine silver staining of lung epithelial cells with phosphate buffered saline (Veh) or *C. albicans* (MOI 1) co-culture. (F) Relative cytotoxicity of lung epithelial cells co-cultured with a yeast-locked (TNRG1) *C. albicans* strain overexpressing the hyphal repressor gene *Nrg1* compared to isogenic control (TT21). (G) Human lung alveolar epithelial cell cytotoxicity as measured by lactate dehydrogenase release with *C. albicans* co-culture at differing multiplicities of infection (MOI). (H-I) Human lung alveolar epithelial cells were grown on transwells at air liquid interface and transepithelial resistance (TEER) was measured at 0 and 24 hours. (H) TEER measurements for *C. albicans* (SC5314) at varying multiplicity of infections compared to vehicle control and (I) yeast-locked (TNRG1) and wild-type (TT21) strains compared to vehicle control. Cytotoxicity results represent three or four independent trials performed in quadruplicate. Values are shown as the median of each trial with interquartile range. TEER values represent individual transwells reported as the ratio of an average of three measurements at 24 hours compared to baseline after blank subtraction and correction for transwell surface area across one or two independent trials.

### LRT Candida colonization is associated with endpoints of lung injury in acute respiratory failure

To examine the clinical relevance of LRT *Candida* colonization on lung injury endpoints, we analyzed clinically available data from a prospective cohort of critically ill patients with acute respiratory failure (ARF), who were intubated and mechanically ventilated in intensive care units at UPMC hospitals (24, 25). To define the exposure of LRT *Candida* colonization, we included 515 patients with ARF who had an available respiratory culture (BAL or endotracheal aspirate-ETA) obtained for clinical purposes within two days of enrollment. Next, we classified the 400 patients with evidence of any microbial growth on cultures, who were stratified into those with bacterial growth (n=324) versus those with yeast growth in cultures (n=76, Table S1). In our institution, yeast growth in respiratory cultures is routinely reported as “yeast not otherwise specified” to indicate *Candida* spp. colonization and not invasive fungal infection. Patients with bacterial versus yeast growth had similar distribution of demographic and baseline clinical variables. However, patients with yeast growth had worse respiratory mechanics, with higher driving and plateau pressure on mechanical ventilation (Figure 6A), fewer ventilatory free days (VFDs) by day 28 (Figure 6B), and higher proportion of detection of neutrophils on Gram stain (88.2%) compared to patients with bacterial growth (62.3%, p<0.001, Figure 6C).

**Figure 6:**
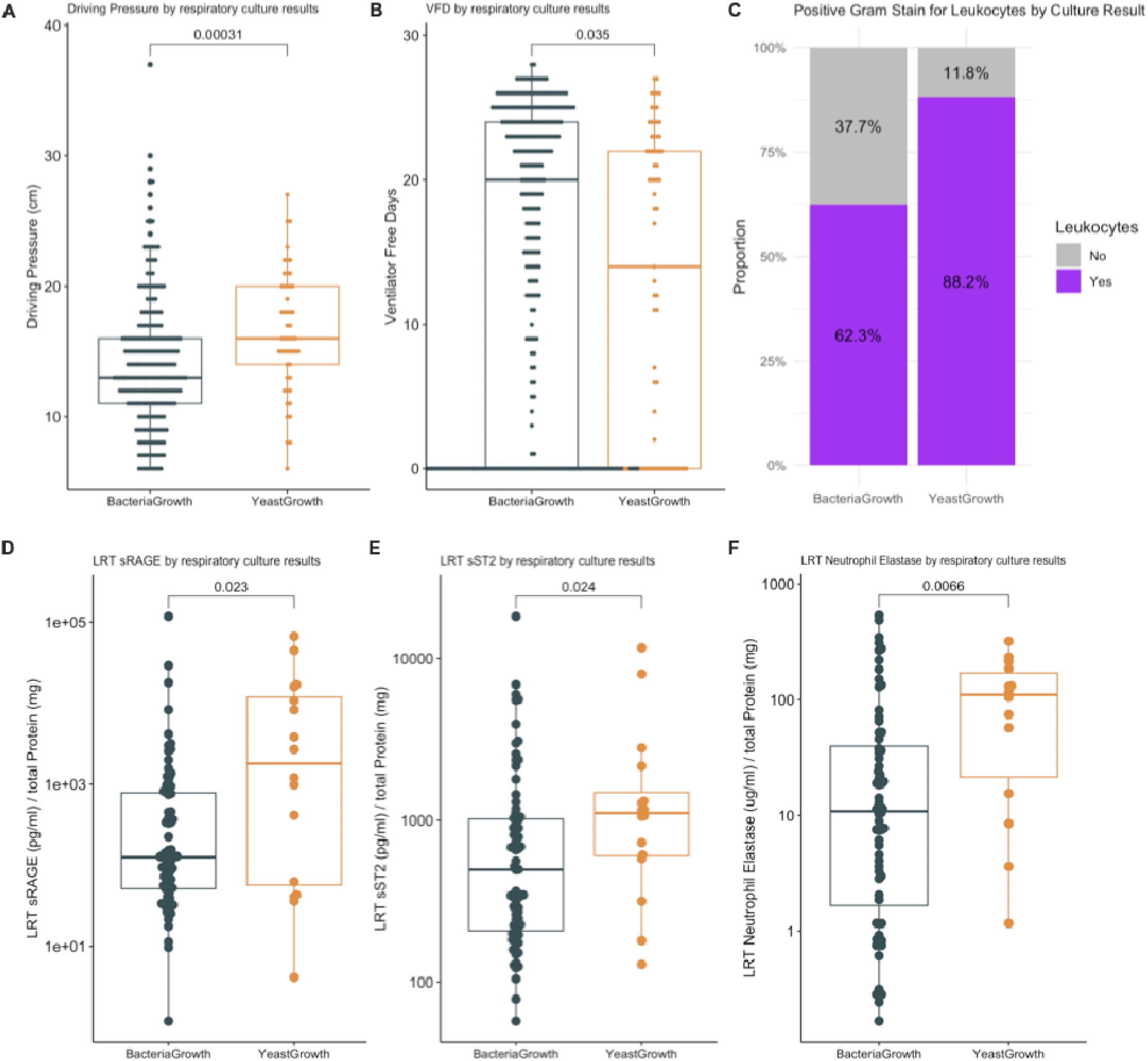
Critically ill patients with lower respiratory tract *Candida* demonstrate worse endpoints of respiratory mechanics, clinical outcomes and lower respiratory track biomarkers of epithelial injury and neutrophil inflammation compared with patients with no evidence of Candida spp in respiratory cultures: Results represent analysis of clinically available data from a prospective cohort of 865 critically ill patients, 515 of which had ARF and an available respiratory culture. Patients were further separated into those with bacterial growth (n=324) or yeast growth (n=76). Comparison of respiratory mechanics represented by driving pressure (A) and clinical endpoints represented by ventilator free days (B) between patients with *Candida* versus bacterial only growth. Proportion of patients with positive gram staining for neutrophils (C) between *Candida* and bacterial only growth. Luminex panel results from endotracheal aspirate supernatant demonstrating biomarkers of epithelial injury including soluble receptor for advanced glycation end products (D), the epithelial alarmin soluble suppressor of tumorigenicity-2 (E), and neutrophil elastase (F) between patients with *Candida* versus bacterial growth.

In a subset of 95 patients with available LRT biospecimens obtained for research purposes upon enrollment, we measured host response biomarkers in ETA supernatants with custom-made Luminex panels (26, 27). Notably, patients with yeast growth had higher ETA levels of the epithelial injury biomarker soluble receptor of advanced glycation end-products (sRAGE), the epithelial alarmin soluble suppressor of tumorigenicity-2 (sST2), and neutrophil elastase (all p<0.05) compared to specimens from patients with bacterial growth (Figure 6D-F). Thus, our clinical observations provided further validation for the associations between the presence of *Candida* in the LRT with biomarkers of lung epithelial injury (sRAGE and sST2), neutrophil presence in the LRT (gram stain leukocytes and neutrophil elastase levels in ETA supernatants), as well as worse respiratory mechanics (driving pressure) and clinical endpoints (VFDs).

## Discussion

In our study, we found that *C. albicans* potentiated LPS-induced lung injury and was itself sufficient to produce early lung injury, characterized by air-blood barrier disruption and neutrophil recruitment into the airspaces. While neutrophils were indispensable for preventing systemic dissemination and mortality following IT *C. albicans*, they were not required for the development of severe lung injury. We demonstrated that *C. albicans* produces direct epithelial cytotoxicity and barrier disruption *in vitro*, both in murine and human lung epithelial cells. Importantly, these findings were validated in critically-ill humans, where higher levels of epithelial injury biomarkers and neutrophil activity in LRT secretions were observed in patients with respiratory cultures positive for *Candida.* These patients also experienced worse clinical outcomes, including fewer ventilatory free days, compared to those with bacterial growth in LRT cultures.

The clinical impact of *Candida spp.* colonization in the LRT of critically ill patients, particularly those on mechanical ventilation, remains unclear (28). Despite frequent recovery of *Candida* in LRT cultures, there has been limited histopathological evidence of *Candida* invading lung tissue, with autopsy studies revealing rare or absent occurrence of pulmonary Candidiasis (0-8% of patients) (29–31). Clinical evidence supporting the efficacy of systemic or localized antifungal treatment for *Candida* spp. colonization is sparse. Two retrospective cohort studies demonstrated that nebulized amphotericin B was effective for *Candida* spp. eradication. (32, 33). However, the *Candida* eradication was associated with measurable clinical benefit in one of these two studies (32), and thus the available evidence cannot justify routine use of nebulized amphotericin B. The CANTREAT pilot trial, which evaluated intravenously administered antifungal therapy in 60 patients with LRT *Candida* colonization at the time of VAP diagnosis, found no significant differences in mortality (8). Notably, CANTREAT was underpowered and investigated antifungal treatment concurrent with antibiotics at the time of VAP diagnosis, which may have been too late in the disease progression to mitigate potential *Candida*-related harm. Therefore, current consensus is that *Candida* spp. in the LRT do not represent an invasive pathogen, and detection of *Candida* spp. in LRT specimen cultures is deemed by many clinicians as an inactionable finding.

Given that *Candida* spp. in the LRT are not routinely treated with antifungals, we have accumulating observational evidence examining the natural history of *Candida* spp. colonization. Both culture-based and culture-independent methods with DNA sequencing of LRT specimens have shown consistent associations between *Candida* spp. detection and adverse clinical outcomes, including prolonged mechanical ventilation and hospitalization, increased risk of VAP, and higher mortality (4–7). These observations have challenged the prevailing assumption that *Candida* colonization in the LRT is harmless (28), and suggest that it may, in fact, contribute to lung injury in ARF.

In our study, we sought to unravel the complex host-pathogen interaction between *C. albicans* and the lung environment using *in vivo* and *in vitro* models. We employed a sterile model of LPS-induced lung injury to control for context-dependent interactions between *C. albicans* and other bacterial organisms. Our experiments revealed that mice treated with *C. albicans* (CAN6LPS) showed increased susceptibility to LPS-induced lung injury compared to vehicle-treated controls (VehLPS). The CAN6LPS mice exhibited higher normalized lung weights, increased BAL total protein, and elevated IgM levels. Histological analysis of CAN6LPS demonstrated neutrophilic infiltration and tissue injury. Taken together, these results are consistent with the American Thoracic Society definitions of experimental acute lung injury (34, 35).

Given the increased lung injury with *C. albicans* pre-exposure, we sought to explore the early effects of *Candida* on lung injury in our model. Notably, the intratracheal instillation of *Candida* alone resulted in significant body weight loss and elevated markers of lung barrier disruption and epithelial injury. *C. albicans* instillation also led to neutrophil influx into the airspaces. Transcriptomic analyses of non-lavaged lung tissue on day 1 revealed gene expression markers of neutrophil degranulation and immune activation, underscoring the inflammatory response triggered by *Candida*.

The importance of neutrophils for both murine and human host defense against invasive fungal infections, including candidemia, is well established (20). Few studies have examined the role of neutrophils in *C. albicans*-induced pulmonary infection. In a fungal septicemia model, *C. albicans* induced complement-mediated neutrophil clustering leading to capillaritis, pulmonary hemorrhage, and hypoxemia (36). Thus, we hypothesized that neutrophils mediated the lung damage we observed with intratracheal *Candida* administration. In our neutrophil ablation model (PMN^DTR^), neutropenic mice were highly susceptible to intratracheal *C. albicans*, with 50% mortality by day 2 and high fungal burden in lung tissue and extra-pulmonary dissemination to the kidneys. Despite reduced neutrophil airspace recruitment, the PMN^DTR^ mice exhibited significantly elevated markers of lung injury compared to wild-type mice, suggesting that *Candida* can also induce lung injury through mechanisms independent of neutrophil-mediated inflammation.

Our findings from both wild-type and genetically-modified murine models suggest that *C. albicans* may play a dual role in lung injury, depending on host immune status. In immunocompetent hosts, neutrophils appear crucial for mediating the lung damage by *Candida*, as their recruitment to the lungs in response to colonization triggers an inflammatory response aimed at controlling the pathogen. However, this protective response can paradoxically exacerbate tissue damage, promoting alveolar-capillary barrier disruption and neutrophil-mediated cytotoxicity. Thus, our findings support a mechanism of *Candida*-related lung injury even in the absence of overt fungal invasion in immunocompetent hosts. Conversely, in the neutropenic model, *C. albicans* caused substantial epithelial damage with unchecked fungal proliferation. Thus, in the immunocompromised host by neutropenia, the role of *Candida* appears to shift from an inflammatory instigator to a direct pathogen capable of causing substantial epithelial damage. We then further traced such pathogenicity by examining candidate virulence genes for direct *Candida*-induced epithelial damage.

To further investigate the mechanisms of *Candida*-induced lung injury, we employed *in vitro* cell culture models. Co-culture of *C. albicans* with murine lung epithelial cells led to dose-dependent cytotoxicity, which was significantly reduced when *Candida* was heat-killed prior to administration. Transcriptomic analysis of *C. albicans*-treated lung epithelial cells demonstrated signatures of NFκB and MAPK activation, similar to prior reports (21), including the hyphae-dependent *Fos*/*Dusp1* pathway. Indeed, *C. albicans*’ ability to transition between yeast and hyphal forms is a key factor in its pathogenicity (22, 37–39). We identified the hyphal form as particularly important in causing lung epithelial damage. Using a yeast-locked mutant strain (TNRG1), we observed decreased cytotoxicity compared to isogenic controls, highlighting the role of hyphal transition in epithelial injury. We reproduced the dose-dependent cytotoxicity phenotype of *C. albicans* in a human lung epithelial cell line. Additionally, we found that *C. albicans* produced lung epithelial barrier disruption in an *in vitro* air-liquid interface culture, which was attenuated with the yeast-locked (TNRG1) mutant strain, further emphasizing the importance of hyphae for pathogenicity.

Our human cohort data further support the relevance of LRT *C. albicans* in clinical settings. Patients with yeast detected in clinical LRT cultures exhibited worse lung injury markers, including elevated levels of epithelial injury biomarkers (sRAGE and sST2), neutrophil elastase, and impaired respiratory mechanics. These findings align with our murine model results, suggesting that LRT *Candida* spp. presence is not a benign phenomenon but rather a contributor to lung injury and inflammation in critically ill patients.

Our study has several limitations. Development of a sustained colonization model in mice is difficult, as our dose-finding experiments showed that all intratracheal inoculums of *C. albicans* had been cleared from the lung by day 4. Sustained *C. albicans* colonization in the LRT in mice would have required repeated administrations, which may have caused more serious injury than the expected transient effects seen in immunocompetent hosts. Here, we focus on day 2 as a surrogate for colonization given consistent recovery of *C. albicans* from the LRT at the 10^6^ inoculums, but we do not know if this directly mirrors human disease progression. Additionally, we show that neutrophils, while critical for curtailing disease severity, are not required to observe lung injury biomarkers. However, other immune or inflammatory cell types may contribute to lung injury in the presence of *C. albicans,* which were outside the scope of our investigation. Finally, the human observational data are subject to confounding by indication, given that availability of LRT cultures depended on clinical indications as deemed appropriate by the treating physicians.

Our study provides compelling evidence that *C. albicans* contributes to lung injury through hyphae-mediated epithelial cytotoxicity and barrier disruption, and that neutrophils, while critical in host defense, are not the sole mediators of *Candida*-associated lung damage. These findings add a new dimension to the understanding of *Candida* colonization in critically ill patients, suggesting that it may be a modifiable risk factor that has long been overlooked. Further research into the molecular and immunological mechanisms underlying *Candida*-induced lung injury could identify novel therapeutic targets and better define the patient populations that would benefit from targeted interventions.

## Materials and Methods

### Animals

C57BL6 mice were purchased from Taconic Biosciences Inc. MRP8-Cre^+^ (B6.Cg-Tg(S100A8-cre,– EGFP)1Ilw/J, stock no. 021614, Jackson Laboratory) mice were crossed with ROSA26iDTR (C57BL/6– Gt(ROSA)26Sortm1(HBEGF)Awai/J, stock no. 007900, Jackson Laboratory) mice to generate a DTR-mediated conditional neutrophil ablation model (PMN^DTR^). The mice generated from crossing MRP8-Cre^-^ with ROSA26iDTR mice were used as littermate controls (PMN^WT^). Eight-to fifteen-week-old male and female mice were used for experiments. All mice were housed in specific pathogen free conditions under the supervision of the Division of Laboratory Animal Resources at the University of Pittsburgh, and experiments were conducted following NIH guidelines under protocols approved by the University of Pittsburgh Institutional Animal Care and Use Committee.

### Two-hit acute lung injury model

*C. albicans* (SC5314) was grown in Yeast extract-peptone-dextrose (YPD) broth at 30°C for 18h in an incubator shaker. On day 0, we intratracheally inoculated mice with 50μ containing 1×10^5^ or 1×10^6^ of *C. albicans* or with 50μl of control phosphate-buffered saline (PBS) solution (Vehicle). On day 2, we intratracheally inoculated mice with either LPS (5mg/kg) to induce acute lung injury with 50μl PBS (Vehicle). The mice were euthanized on Day 4 and underwent blood sampling by venipuncture of the inferior vena cava, bronchoalveolar lavage (BAL) sampling of the right lung, and harvesting of the left lung, kidneys, liver, spleen, stomach, ileus and colon for further analysis.

### Acute lung injury model due to intratracheal *C. albicans*

*C. albicans* (SC5314) was grown in Yeast extract-peptone-dextrose (YPD) broth at 30°C for 18h in an incubator shaker. On day 0, we intratracheally inoculated mice with 50μl of solution containing 1×10^6^ of *C. albicans* or with 50μl of control phosphate-buffered saline (PBS) solution (Vehicle). Mice were euthanized on Day 1 or 2 and underwent blood sampling by venipuncture of the inferior vena cava, bronchoalveolar lavage (BAL) sampling of the right lung, and harvesting of the left lung, kidneys, liver, spleen, stomach, ileus and colon for further analysis.

### Bronchoalveolar lavage (BAL)

BAL was centrifuged at 2,000g for 10 min at 4°C and supernatant was stored at –80°C until further analysis. The pellet was resuspended with PBS after red blood cell lysis by ACK Lysing Buffer (Thermo Fisher Scientific). The number of cells in the BAL was counted with a hemocytometer. The percentage of neutrophils in the BAL was analyzed by flow cytometry. Staining of neutrophils was performed with CD45-PerCP-Cy5.5 (BD Pharmingen), Ly6G-FITC (Invitrogen), Ly6G-eFluor450 (Invitrogen), and CD11b-AlexaFluor700 (BioLegend) antibodies. Dead cells were excluded using Ghost dye violet 510 (Tonbo Bioscience).

### *C. albicans* tissue cultures

We homogenized tissues with the GentleMACS dissociator (Miltenyi Biotec) and then plated homogenate suspensions on YPD agar plates after serial dilution. Plasma from the inferior vena cava was collected into K2 EDTA tubes (BD) and mixed briefly prior to serial dilution. We incubated YPD agar plates for 24h in 30°C incubator and measured fungal burden as colony forming units (CFUs) per gram of tissue or per milliliter.

### Lung histology

In a dedicated experiment for lung histology, we harvested lung tissues following lung perfusion and agar inflation. We fixed tissues with 10% formalin and then processed them for sectioning and staining with hematoxylin-eosin (H&E). We selected 12 random fields from each stained tissue section and captured images with a Nikon microscope. A blinded independent reviewer examined all images on 20x magnification and performed lung injury histology scoring according to the American Thoracic Society guidelines (35).

### Biomarker measurements

We measured murine cytokines and chemokines with a 23-plex Luminex panel (Bio-Plex Pro Mouse Cytokine, Bio-Rad, Hercules, CA) according to manufacturer’s instructions in BAL supernatant (1.5-fold dilution), plasma (5-fold dilution), and lung and kidney tissue homogenates (4-fold dilution). In BAL fluid supernatants, we measured total protein with a BCA assay (undiluted samples), IgM (in both undiluted and 40-fold diluted samples) with ELISA (Invitrogen, Thermo Fisher Scientific), and RAGE (in 2-fold diluted samples) with ELISA (Duoset, R&D Systems).

### Bulk RNA sequencing of mouse lung

Snap-frozen left lungs collected at day 1 from vehicle-(n=4) or *C. albicans*-treated (n=4) mice were used for RNA isolation. Mouse lungs were homogenized in TRIzol (Invitrogen), and RNA was isolated per manufacturer’s instructions. RNA clean-up and purification were performed using the RNeasy Mini Kit (Qiagen) per manufacturer’s instructions. Library preparation, quality control, and paired end sequencing were performed by Novogene. FastQ files were trimmed and aligned to generate a feature count matrix using the Center for Research Computing HTC shell. Differential gene expression, overrepresentation, and gene set enrichment analyses were performed in RStudio (version 4.3.1) using the DESeq2, clusterProfiler, and fgsea packages, respectively.

### *In vitro* cell death assays

Mouse lung epithelial (MLE-12, ATCC) cells or human alveolar epitheial cells (CI-huArlo, InSCREENeX) were plated in a 96-well plate at 2 × 10^4^ cells per well 16 hours prior to *C. albicans* exposure. *C. albicans* (SC5314) was grown to saturation in YPD broth in an incubator shaker for 18 hours at 30°C and 250 RPM. The *C. albicans* was then centrifuged and washed three times with phosphate buffered saline (PBS) and counted using a hematocytometer. To obtain different multiplicity of infections (MOI), the *C. albicans* was diluted with PBS. For the heat-killed experiments, *C. albicans* was incubated at 90°C for 30 minutes after dilution to the appropriate MOI. For yeast mutant experiments, TT21 (wild type) and TNRG1 (yeast-lock) strains were used instead of SC5314. The diluted *C. albicans* solutions were combined with fresh, low-serum media (RPMI 1640, 2% fetal bovine serum for MLE-12 and huAEC medium for CI-huArlo) and then added to the appropriate wells. The plate was placed in a bacterial incubator at 37°C with 5% CO2 for 6 hours. The supernatant was transferred to a new 96 well plate and centrifuged at 125 x g for 5 minutes to pellet cells and debris. Cell death was measured via LDH release using the LDH-Glo Cytotoxicity Assay (Promega) per the manufacturer’s instructions. The percent cytotoxicity was calculated as previously described (40). Of note, these calculations include a subtraction of background *C. albicans* LDH release and use of 2% Triton X as a positive control. If the calculated cytotoxicity was below background, the value is reported as 0. We performed each experiment in quadruplicate and repeated across three or four independent trials.

### Transepithelial resistance (TEER) measurements

Human alveolar epithelial cells (CI-huArlo, InSCREENeX) were plated onto a 12-well transwell (Corning) after pre-coating per manufacturer’s instructions. The cells were grown at 37°C and 5% CO2 in submerged culture until confluent and then the apical media was removed to create an air-liquid interface. The cells were incubated for an additional two weeks to allow for tight barrier development. On the day of the experiment, transwells were moved into a new, sterile 12-well plate and phosphate buffered saline (PBS) was added to the apical and basolateral surfaces. Transepithelial resistance was measured using the Millicell-ERS Voltohmmeter (Millipore Sigma) per the manufacturer’s instructions. The transwells were then replaced into the 12-well plate, fresh media was added to the basolateral side, and *C. albicans* or PBS (vehicle) in CI-huArlo culture media was added to the apical surface at a multiplicity of infection (MOI) of 1 or 5. The transwells were incubated at 37°C and 5% CO2 for 24 hours. The apical solution was aspirated and the TEER was measured again as described above. Three values were measured per well at both baseline and 24 hours and averaged. Values were subtracted from a PBS only measurement and adjusted for transwell surface area. Values represent individual transwells across 1 or 2 independent experiments.

### Bulk RNA sequencing of murine lung epithelial cells

MLE-12 cells were plated in a 12-well plate at 0.5 × 10^6^ cells/well 16 hours prior to treatment. *C. albicans* (SC5314) was prepared as previously described to achieve an MOI of 0.5. *C. albicans* were heat killed (HKCA) at 90°C for 30 minutes. Cells were then treated with either phosphate buffered saline control (n=4), *C. albicans* (n=4), or HKCA (n=4) in fresh, low serum media (RPMI 1640, 2% FBS). Cells were incubated at 37°C and 5% CO2 for 4 hours. The treatment media was aspirated, and the cells were washed once with fresh RPMI. Buffer RLT (Qiagen) with 1% v/v β added to each well. The cells were incubated on ice for 5 minutes. Cell lysates were collected immediately and passed through a QIAShredder tube (Qiagen) per manufacturer’s instructions. RNA purification, sequencing, and down-stream analyses were performed as described in the lung RNA sequencing methods section.

### Murine lung epithelial cell microscopy

MLE-12 cells were plated onto sterile, ethanol-treated coverslips in a 24-well plate at 0.5 × 10^6^ cells/well 16 hours prior to treatment. *C. albicans* (SC5314) was prepared as previously described to achieve an MOI of 1. Cells were treated with either phosphate buffered saline (Vehicle) or *C. albicans* (MOI 1). The plate was incubated at 37°C and 5% CO2 for 6 hours. The cells were washed 3 times with PBS and fixed with 2% paraformaldehyde (PFA) in PBS for 15 minutes at room temperature. The PFA was removed and replaced with fresh PBS. The coverslips were stored at 4C until use. Coverslips were stained using Grocott’s methenamine silver stain (Newcomer Supply) per the manufacturer’s instructions. Stained coverslips were imaged on an Olympus IX73 widefield microscope using a 60X Plan Apochromat (NA 1.42) lens.

### Human cohort data

Our research complies with all ethical regulations in accordance with the Declaration of Helsinki and as directed by the University of Pittsburgh Institutional Review Board (IRB) (protocol STUDY19050099). Following admission to the ICU at UPMC (Pittsburgh, PA, USA) and obtaining informed consent from patients or their legally authorized representatives, we collected baseline research biospecimens within 72hrs from intubation. We collected endotracheal aspirates (ETA) for host biomarker measurements. We obtained clinical data directly from the electronic medical record. We captured biological sex and race as recorded in the medical record. A consensus committee reviewed clinical and radiographic data and performed retrospective classifications of the etiology and severity of acute respiratory failure without knowledge of research assay data. We retrospectively classified subjects as having ARDS per established criteria (Berlin definition), being at risk for ARDS because of the presence of direct (pneumonia or aspiration) or indirect (e.g., extrapulmonary sepsis or acute pancreatitis) lung-injury risk factors although lacking ARDS diagnostic criteria, having acute respiratory failure without risk factors for ARDS, or having acute-on-chronic respiratory failure.

We followed patients prospectively for cumulative mortality and ventilator-free days (VFDs) at 28 days. We recorded clinical microbiologic culture results obtained within 2 days from participant enrollment to the cohort. We measured biomarkers of tissue injury and inflammation with custom Luminex multi-analyte panels in ETA supernatants, as previously described. We measured total protein in ETA supernatants with a BCA assay.

### Statistical analysis

All data are expressed as medians ± interquartile ranges (IQR), unless otherwise specified. We performed univariate analyses using ANOVA, Mann-Whitney, and Kruskal-Wallis test with GraphPad Prism 10 program. For tissue transcriptomics, we examined for differentially abundant genes adjusted for multiple comparisons with the Benjamini-Hochberg method and the *DESeq2* package in R, whereas for differentially expressed pathways, we used the *fgsea* package for Gene Set Enrichment Analysis (GSEA).

## Figure legends

**Figure S1:**
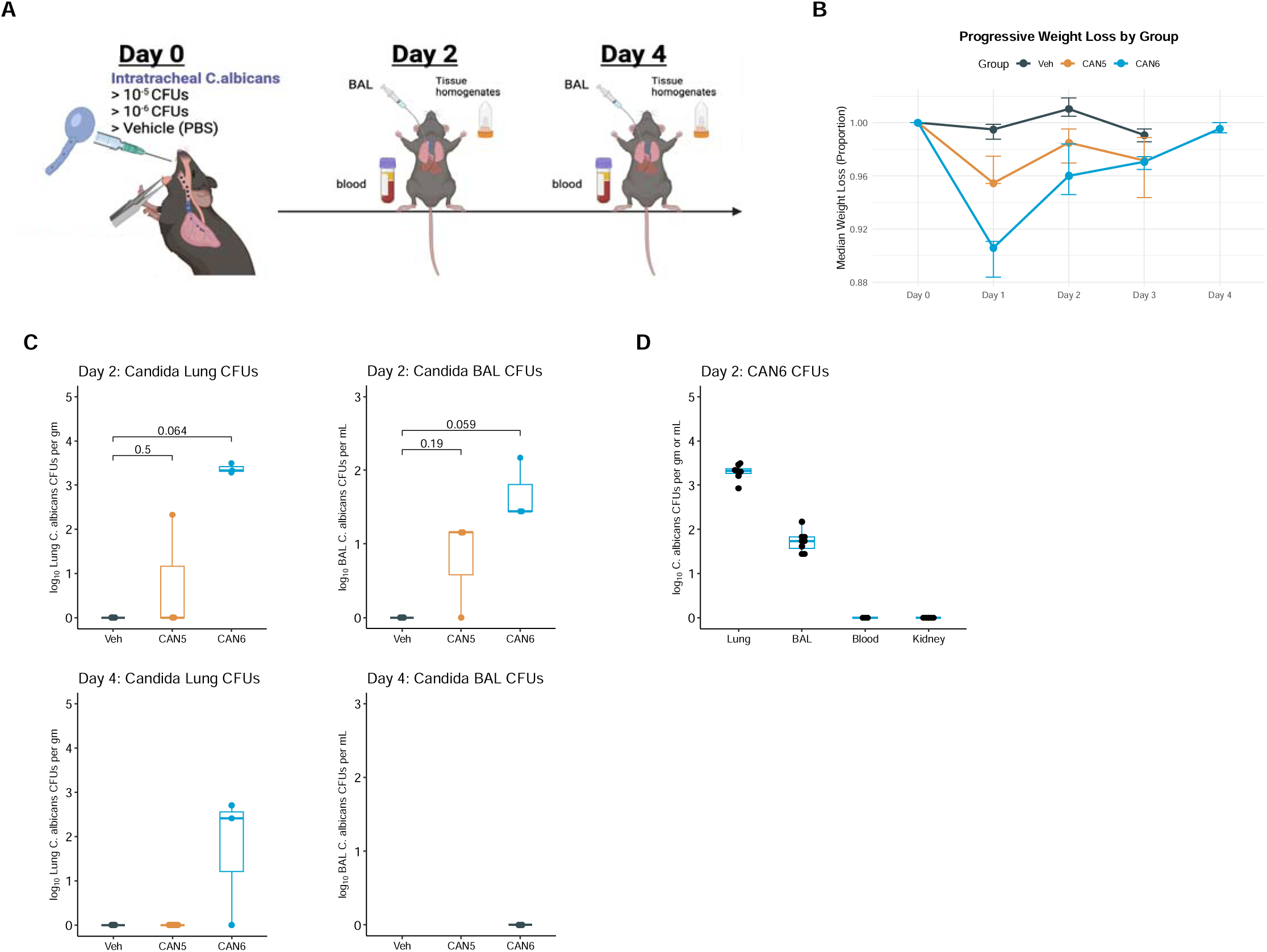
Validation of an intratracheal *C. albicans* colonization mouse model. (A) Mice received 10^5^ or 10^6^ colony forming units (CFUs) (ITCAN5 or ITCAN6, respectively) of *C. albicans* or phosphate-buffered saline (Veh) on day 0 and were scarified on day 2 or 4. (B) Median body weight loss for each experimental condition. (C) Log-transformed *C. albicans* colony forming units per gram of lung homogenate or milliliter of bronchoalveolar lavage on day 2 or 4 from the right lung of scarified mice. (D) Log-transformed C. albicans colony forming units per gram of kidney and lung and per milliliter of BAL and blood on day 2.

**Table S1.** Clinical characteristics of patients with acute respiratory failure and bacterial growth (n=324) or yeast growth (n=76) in lower respiratory tract specimens. Data are presented as median (interquartile range) or n (%) as appropriate. Abbreviations: ARDS: acute respiratory distress syndrome, COPD: chronic obstructive pulmonary disease, mSOFA: modified sequential organ failure assessment [does not include the neurologic component].

|  | Bacterial growth | Fungal Growth | p-value |
| --- | --- | --- | --- |
| N | 324 | 76 |  |
| Demographics |  |  |  |
| Age, years | 58.9 [46.7, 68.1] | 54.4 [42.2, 67.5] | 0.06 |
| Sex, male | 192 (59.3) | 44 (57.9) | 0.93 |
| Race |  |  |  |
| Caucasian | 292 (90.1) | 68 (89.5) |  |
| African American | 26 ( 8.0) | 7 ( 9.2) |  |
| Body mass index | 29.5 [25.1, 36.0] | 29.3 [25.5, 36.3] | 0.9 |
| Comorbid Conditions |  |  |  |
| Congestive heart failure | 37 (11.4) | 12 (15.8) | 0.39 |
| Diabetes mellitus | 118 (36.4) | 19 (25.0) | 0.08 |
| COPD | 75 (23.1) | 12 (15.8) | 0.21 |
| Alcohol use | 43 (13.3) | 7 ( 9.2) | 0.44 |
| Chronic kidney disease | 51 (15.7) | 17 (22.4) | 0.22 |
| Active malignancy | 16 ( 4.9) | 4 ( 5.3) | 1 |
| Immunosuppression | 68 (21.0) | 26 (34.2) | 0.02 |
| Chronic liver disease | 27 ( 8.3) | 8 (10.5) | 0.7 |
| Pulmonary fibrosis | 16 ( 4.9) | 6 ( 7.9) | 0.46 |
| Vital signs |  |  |  |
| Temperature (°C) | 36.9 [36.2, 37.6] | 36.8 [36.2, 37.3] | 0.23 |
| Heart rate (beats per minute) | 92.0 [79.0, 104.0] | 96.0 [80.5, 107.5] | 0.12 |
| Respiratory rate (breaths per minute) | 20.0 [17.0, 25.0] | 22.0 [17.0, 26.2] | 0.36 |
| Systolic blood pressure (mmHg) | 117.0 [104.0, 133.0] | 115.5 [103.5, 128.2] | 0.3 |
| Oxygen saturation (%) | 97.0 [95.0, 99.0] | 97.0 [94.0, 99.0] | 0.48 |
| Ventilatory characteristics |  |  |  |
| Tidal volume (mL/kg) | 6.7 [6.0, 7.6] | 6.7 [5.9, 7.4] | 0.55 |
| Minute ventilation (L/min) | 9.6 [7.8, 11.5] | 9.6 [7.9, 11.9] | 0.9 |
| Positive end expiratory pressure (cm H2O) | 8.0 [5.0, 10.0] | 10.0 [5.0, 10.0] | 0.04 |
| Peak inspiratory pressure (cm H2O) | 25.0 [20.0, 32.0] | 29.0 [25.0, 33.0] | <0.01 |
| Plateau pressure (cm H2O) | 21.0 [17.0, 26.0] | 25.0 [21.0, 28.0] | <0.01 |
| Worst PaO2/FiO2 ratio | 154.0 [95.0, 205.0] | 137.0 [82.0, 197.5] | 0.17 |
| Clinical laboratory values |  |  |  |
| White blood cells x 10^9/L | 12.8 [8.7, 18.0] | 12.2 [9.3, 16.3] | 0.69 |
| Hemoglobin, g/L | 10.6 [9.3, 12.3] | 9.9 [8.6, 11.1] | <0.01 |
| **Platelets x 10<sup>9</sup>/L** | 192.0 [139.2, 253.5] | 183.0 [129.5, 252.5] | 0.49 |
| **Creatinine, mg/dL** | 1.2 [0.8, 1.9] | 1.4 [0.8, 2.4] | 0.28 |
| **Bicarbonate** | 24.0 [21.0, 28.0] | 24.0 [22.0, 28.5] | 0.76 |
| **Arterial pH** | 7.4 [7.3, 7.4] | 7.4 [7.3, 7.4] | 0.32 |
| **Arterial paCO2** | 44.0 [38.0, 52.0] | 47.0 [40.0, 56.0] | 0.34 |
| **Severity of illness** |  |  |  |
| **mSOFA** | 7.0 [5.0, 9.0] | 7.0 [5.0, 9.0] | 0.87 |
| **Pneumonia** | 185 (58.0) | 46 (62.2) | 0.6 |
| **Non-pulmonary sepsis** | 47 (14.7) | 11 (14.9) | 1 |
| **Aspiration** | 57 (17.9) | 7 ( 9.5) | 0.11 |
| **Subphenotypes** |  |  |  |
| **Hyperinflammatory (Drohan Model)** | 60.8 (19.7) | 23 (34.3) | 0.01 |
| **Hyperinflammatory (Sinha Model)** | 52 (16.8) | 16 (23.9) | 0.24 |
| **Clinical outcomes** |  |  |  |
| **Ventilator-free days** | 20.0 [0.0, 24.0] | 14.0 [0.0, 22.0] | 0.04 |
| **30-day Mortality** | 82 (25.3) | 21 (27.6) | 0.79 |
| **90-day Mortality** | 101 (31.2) | 25 (32.9) | 0.88 |

## Data Availability

All data produced in the present study are available upon reasonable request to the authors

